# Characterisation of multimodal network organisation after focal prefrontal lesions in humans

**DOI:** 10.1101/2020.12.09.20216903

**Authors:** MP Noonan, MR Geddes, RB Mars, LK Fellows

## Abstract

Lesion research in humans and non-human primates classically maps the behavioral effects of focal damage to the directly-injured brain region. However, given the interconnectedness of the brain, it has long been known that such damage can also have distant effects. Modern imaging methods provide new ways to assess those effects. Further, triangulating across these methods in a lesion model may shed light on the biological basis of structural and functional networks in the healthy brain. We characterised network organization assessed with multiple MRI imaging modalities in 13 patients with chronic focal damage affecting either superior or inferior frontal gyrus (SFG, IFG) and 18 demographically-matched healthy Controls. We first defined structural and functional network parameters for the two frontal regions-of-interest in healthy Controls, and then used voxel-based morphology (VBM) and tract-based spatial statistics (TBSS) analyses to investigate structural grey matter (GM) and white matter (WM) differences between patients and Controls. The functional and structural networks defined in healthy participants were then used to constrain interpretation of the whole brain network effects in patients. Finally, we applied dual regression to examine the differences in functional coupling to large-scale resting state networks (RSNs), focusing on the RSNs which most overlapped structurally with the lesion sites. Overall, the results show that lesions are associated with widespread within-network GM loss at sites distal from the lesion, yet leave WM and RSNs relatively preserved. Lesions to either prefrontal region had a very similar impact on structural networks, but SFG lesions had larger impact on RSNs than did IFG lesions. The findings provide evidence for causal contributions of specific prefrontal regions to structural and functional brain networks in humans, relevant to interpreting connectomic findings in studies of healthy people or those with psychiatric illnesses.

## Introduction

Although it has long been known that focal brain damage can have distant effects, the neuroimaging tools needed to characterize such effects in living humans have only recently become available. Diaschisis (from Greek, meaning “shocked throughout”) refers to a loss of neuronal activity due to acute loss of afferents from a lesioned brain region, and was first described by von Monakow (1914). The rapidly expanding field of network neuroscience has defined a variety of structural and functional networks and linked variation in these networks to individual differences and pathological conditions (Honey & Sporns, 2008; van den Heuvel & Sporns, 2013). However, the biological basis of these networks has been inferred largely from correlational evidence from healthy people and remains a matter of debate. Interpreting the findings in this evolving field is further complicated by the plethora of network analysis methods.

A more robust understanding of the anatomical and physiological basis of these networks is needed. Methods that perturb networks, such as lesions or other loss-of-function approaches, are likely to be particularly useful. Loss-of-function methods provide tests of the necessary contribution of a region to one or more networks. Loss-of-function effects on networks defined with different MRI methods in the same individual can address whether these distinct network measures reflect a common underlying biology (Reid *et al*., 2019). Using this approach, the current study addresses two outstanding questions: First, what is the impact of a lesion on brain networks identified through structural or resting state functional neuroimaging? Second, is the impact of a lesion on brain organization dependent on the lesion location?

Just a few studies to date have examined the effects of focal brain damage on networks. Most have addressed distant effects of an acute lesion (predominantly ischemic stroke in the sensorimotor system (Grefkes *et al*., 2008; Wang *et al*., 2010; Park *et al*., 2011; Rehme *et al*., 2011; Wang *et al*., 2012; Fan *et al*., 2013; Golestani *et al*., 2013; Abela *et al*., 2015; Siegel *et al*., 2016; Almeida *et al*., 2017), by applying unimodal imaging (typically functional MRI) to assess lesion impact on structural or functional connectivity (Catani & ffytche, 2005). Collectively, the findings suggest that compromised network parameters can be related to clinical symptoms (Carter *et al*., 2010; Gratton *et al*., 2012; Baldassarre *et al*., 2014; Dacosta-Aguayo *et al*., 2014; Dacosta-Aguayo *et al*., 2015; Baldassarre *et al*., 2016; Kuceyeski *et al*., 2016), and that clinical recovery is associated with network normalization (Park *et al*., 2011; Dacosta-Aguayo *et al*., 2014; Siegel *et al*., 2018) or changes in the remaining networks (Gauthier *et al*., 2008).

Unfortunately, the emerging picture is less clear in more anterior brain regions and there is still substantial uncertainy over the extent to which lesions affect inter-network connectivity. On the one hand, Nomura and colleagues (2010) reported that damage to either the cingulo-opercular or frontoparietal networks affected functional connectivity among the other, undamaged network-specific nodes and spared nodes outside the damaged network. By contrast, Eldaief and colleagues (2017) reported that lesions of the anterior mPFC, a prominent default mode network (DMN) node, do not weaken intrinsic within-network functional connectivity among undamaged nodes. Instead, network-specific changes manifested as weaker correlations between whole brain resting state networks (RSNs) including the DMN and attentional and somatomotor networks.

While this research lays important groundwork in understanding the biological basis of these networks, it does not take advantage of the convergent evidence that can be acquired by assessing brain networks using multiple modalities. To our knowledge, there are no studies that examine the effects of focal brain lesions on structural and functional networks imaged in the same people (although see Buch *et al*., 2012, using MEG and DWI; and indirect structural measures in Salvalaggio *et al*., 2020). However, researchers have recognized the potential offered by such multimodal approaches, and have used computational models to simulate the effects of focal lesions on white matter (WM) and functional connectivity networks. Directly relevant to the current study, Alstott and colleagues used an anatomically-informed model of large-scale functional and structural connectivity, derived from diffusion spectral imaging sequences and selectively deleted nodes in different areas of the model brain (Alstott *et al*., 2009). The authors observed different changes in connectivity patterns depending on lesion location and whether they were measuring structural or functional connectivity: For example, the structural integrity of a network, defined by connectivity measures derived from WM tracts, was relatively resilient to node deletion, while node deletion had more pronounced effects on functional connectivity. Even when ‘central’ nodes were targeted, this had minimal impact on structural network integrity until over 15% of nodes were deleted. By contrast lesions that comprised of only 5% of nodes had signifant impact on functional connectivity that depended on lesion location. This model showed that simulated lesions along the cortical midline extensively disrupted functional connectivity, while simulated lesions involving lateral regions only affected local areas of the model brain. For example, simulated lesions involving superior medial PFC strongly reduced functional coupling of many ipsi-lesional regions. By contrast, more lateral simulated lesions such as one centred on pars opercularis, mainly reduced local coupling. Finally, simulated lesions involving visual cortex had little effect on functional coupling of the model brain beyond the immediate vicinity of the lesion. These hypotheses have not been tested empirically.

In the present study we acquired multimodal structural and resting state functional MRI data in the same sample of patients with chronic frontal lobe lesions involving either SFG or right IFG (rIFG). We assessed lesion effects on grey matter (GM), WM and resting state functional connectivity, comparing lesion groups to healthy Controls. First, we ask what, if any, is the impact of a SFG or rIFG lesion on brain networks identified through structural or resting state functional neuroimaging. Based on the simulated lesion results (Alstott *et al*., 2009), we predicted lesions would affect functional connectivity to a greater extent than structural white matter connectivity, and potentially by extention (although not simulated in the model) GM. Second, we asked whether the impact of a lesion on a network depends on the lesion location, testing the general claim that midline lesions produce more widespread disruption: specifically, in this sample, we hypothesized that SFG lesions would be associated with greater network disruption than rIFG lesions.

## Methods

### Participants

Fourteen people with focal lesions involving the frontal lobes were recruited from the Cognitive Neuroscience Research Registry at McGill University. Individuals with prefrontal damage were eligible if they had a focal damage affecting one of the frontal lobe regions of interest: rIFG or SFG. One participant could not complete the MRI due to joint pain, leaving 13 participants as the final sample (8 women; mean age (standard deviation (SD)) = 58 (12.9) y). Age- and education-matched healthy Control participants (*n* = 18, 11 women; mean age (SD) = 51.9 (15.3) y) were recruited through local advertisement in Montreal. They were free from neurological or psychiatric disease and not taking psychoactive medication. Demographic information is reported in Table 1. Controls completed screening tests for mild cognitive impairment and depression. All scored 26 or greater on the Montreal Cognitive Assessment (MoCA) (Nasreddine *et al*., 2005), and less than 12 on the Beck Depression Inventory. Patients completed a more extensive neuropsychological screening battery testing memory, language, attention and executive functions at the time of enrolment in the Registry (Table 2). Lesion groups were compared using independent samples t-tests. Lesions were due to ischemic stroke (*n* = 7), low-grade tumour resection (*n* = 5), fast-growing glioma (*n* = 1). The median time since the lesion occurred was 5 years. Patients with focal frontal lobe damage were recruited through the Cognitive Neuroscience Research Registry at McGill University (Fellows *et al*., 2008). All participants provided written, informed consent prior to participation in the study, in accordance with the Declaration of Helsinki, and were compensated for their time with a nominal fee. Participants were compensated for their time. The study protocol was approved by the Institutional Review Board at McGill University.

**Table 1.** Demographic information

| Group | Age (years) | Education (years) |
| --- | --- | --- |
| SFG | 59.7 (11.7) | 15.1 (2.7) |
| rIFG | 55.8 (15.2) | 13.9 (4.4) |
| Controls | 52.9 (15.3) | 15.8 (3.4) |

**Table 2.** Neuropsychological screening

| Group | BDI | IQ estimate (WASI) | Animal fluency | F-A-S fluency | Backwards digit span |
| --- | --- | --- | --- | --- | --- |
| SFG | 9 (11) | 113 (7)** | 17 (5) | 36 (19) | 5.9 (2.1) |
| rIFG | 13 (6) | 109 (13)* | 16 (4) | 34 (19) | 6.6 (2.3)* |

\* 1 participant missing
\*\* 2 participants missing

### Image Acquisition

All images were acquired on a 1.5T Siemens MR scanner at the McConnell Brain Imaging Centre at the Montreal Neurological Institute, McGill University. Participants laid supine in the scanner and cushions were used to reduce head motion. BOLD fMRI data were acquired using echo planar imaging (EPI) (36 x 4 mm thick axial slices with a base resolution of 64mm, field of view 256 x 256 x 144mm^3^, giving a voxel size of 4 x 4 x 4mm, repetition time = 2.8s, 153 volumes, echo time = 50 ms, and flip angle = 90°). The EPI scanning sequence lasted 7 minutes 20 secs and participants were instructed to keep their eyes closed, think of nothing and stay awake. A T1-weighted anatomical image was acquired for each participant (repetition time = 2800 ms, echo time = 4.12 ms, and flip angle = 15°, giving a voxel size of 1 x 1 x 1 mm). Diffusion MRI (DMRI) data were also acquired from 17 of the same participants described above with the same scanner. A technical issue meant it was not possible to collect the DMRI in the 18^th^ Control participant. DMRI data were acquired using echo planar imaging (75 slices, 2 mm thick axial slices; field of view, 256 x 256 x 150 mm; giving a voxel size of 2 x 2 x 2 mm). Diffusion weighting was isotropically distributed along 99 directions using a B value of 1000 mm^2^. Ten volumes with no diffusion weighting were acquired throughout the acquisition. The total scan time for the DMRI protocol was 20.21 min. Note, the data that support the findings of this study are available from the corresponding author upon reasonable request.

### Preprocessing

Data were analyzed using tools from the FMRIB Software Library (www.fmrib.ox.ac.uk/fsl). All structural and EPI images were converted to nifti and skull stripped with BET; where appropriate this stage was corrected by hand. All brain images are shown in the radiological convention.

### Structural reorganisation after frontal lesions

We examined GM and WM differences between lesion groups and Controls using voxel-based morphology (VBM) and tract-based spatial statistics (TBSS). To quantify these VBM effects and inform our interpretation, we concurrently examined connectivity of the two lesion sites in the healthy Control group, as well as a non-lesioned Control site in the patients. We chose to compare against a control site to remove the possibility of biasing the results if one lesion location had a broader connectivity pattern than another. Specifically, we conducted two additional connectivity analyses in the healthy Control sample: probabilistic tractography and seeded resting state. These analyses are described in Supplementary Methods: *Probabilistic tractography* and *Seeded resting state*.

#### Voxel based morphology

We used voxel-based morphometry (Douaud *et al*., 2007 http://fsl.fmrib.ox.ac.uk/fsl/fslviki/FSLVBM) to identify areas of GM where volume differed by group. The skull stripped T1-weighted structural images were individually segmented into GM, WM and cerebral spinal fluid before being affine-registered to the GM ICBM-152 template using FLIRT (Jenkinson & Smith, 2001) followed by nonlinear registration using FMRIB’s Nonlinear Image Registration Tool (FNIRT) (Andersson *et al*., 2008). A study-specific template was created using six participants from each group so as not to bias the structural template. All native GM images from the whole sample were then non-linearly re-registered and concatenated into a 4D image. The registered partial volume images were then modulated (to correct for local expansion or contraction) by dividing by the Jacobian of the warp field. The modulated segmented images were then smoothed with an isotropic Gaussian kernel with a sigma of 3 mm. Segmentation and registration was confirmed by visual inspection.

The resulting 4D image was then used within two independent GLM analyses. The GLM was identical for each analysis and included factors of the group, sex, age, handedness and number of years of education and was implemented using permutation-based non-parametric testing with Randomise (*n* = 5,000), corrected for multiple comparisons (*p* < 0.05) over a GM mask with threshold free cluster enhancement (tfce) methods (Smith & Nichols, 2009). Each analysis differed only by the group contrasts and the GM inclusion mask. Specifically, Controls and SFG patients were compared in one analysis excluding any SFG lesion damaged voxel. Controls and rIFG patients were compared in one analysis excluding any rIFG lesion damaged voxel. We examined positive and negative group contrasts in each analysis. Because the two lesions affect common networks (Aron *et al*., 2014) we chose to compare each lesion group to healthy controls and not to each other, as the latter method may have caused us to miss effects at the other lesion site.

#### Tract-based spatial statistics

We assessed group differences in WM integrity with the FSL TBSS processing pipeline (Smith *et al*., 2006 http://fsl.fmrib.ox.ac.uk/fsl/fslwiki/TBSS). Specifically, the pre-processed data were subjected to DTIFIT, an analysis step which fits a diffusion tensor model at each voxel in order to generate a 3D fractional anisotropy image for each participant. This image was registered to the FMRIB58FA standard brain before a study specific skeletonised FA template was generated and thresholded at 0.2. All participants’ skeletonized FA images were concatenated into a 4D image.

We focused exclusively on the WM tracts identified in healthy Controls as emanating from the lesion sites and terminating in grey matter that overlapped with the VBM lesion effects (see *Supplementary Methods: Probabilistic tractography)*. The probtrack group tractograms seeded bilaterally at the lesion coordinates in healthy Controls were used as small volume of interest masks to constrain the voxel-wise group statistics that used non-parametric permutation testing (Nichols & Holmes, 2002) with Randomise (Winkler *et al*., 2014). The GLM included factors of Group as well as the confound regressors of age, sex and number of years in education. All reported statistics were found to survive cluster correction for multiple comparisons (*p* < 0.05) with tfce methods (Smith & Nichols, 2009). Again, any lesioned voxel was excluded from the statistical tests. For visualisation and identification of WM tracts identified by the TBSS analysis we ran probabiltic tractography seeded from 216mm^3^ masks placed at the centre of gravity of each cluster. Probablistic tractotracty parameters and details related to the contruction and post-processing of tractograms were identifical to those used in the probablisitic tractography analyses in healthy Controls and are described in *Supplementary Methods: Probabilistic tractography*.

### Functional reorganisation of resting state networks after frontal lesions

#### Dual Regression

Each participant’s individual functional EPI data were first preprocessed using Multivariate Exploratory Linear Optimized Decomposition into Independent Components (MELODIC). Components that were clearly caused by head motion or spikes were removed.

Resting state functional connectivity was assessed using the dual regression technique (Filippini *et al*., 2009 http://fsl.fmrib.ox.ac.uk/fsl/fslwki/DualRegression). This three-step method allows for voxel-wise comparisons of resting state network functional connectivity. We examine the differential contribution of voxels in the brain to large-scale RSNs between patients and Controls and as such our voxel-wise methods are equivalent across GM, WM and functional connectivity. First, all participants’ denoised resting-state functional MRI (rsfMRI) data were collectively motion corrected, spatially smoothed (using a Gaussian kernel of full-width at halfmaximum (FWHM) of 6 mm) and high-pass temporally filtered to 150 s (0.007 Hz). Individual fMRI volumes were registered to the individual’s structural scan and standard space images using FNIRT. Pre-processed functional data containing 154 time points for each Control participant were temporally concatenated across participants to create a single group 4D FMRI data set. Note, no lesioned data were included in this initial ICA decomposition. This concatenated group data set is then decomposed using independent component analysis (ICA). ICA is a data-driven approach used to identify large-scale patterns of functional connectivity in the healthy population of participants. In this analysis, the data set was decomposed into 20 components, in which the model order was estimated using the Laplace approximation to the Bayesian evidence for a probabilistic principal component model.

The second step uses the dual-regression approach to identify, within each participant’s fMRI data set, subject-specific temporal dynamics and associated resting state network (RSN) spatial maps. This step was run on all lesion and Control participants and involves (i) using the full set of group-ICA spatial maps in a linear model fit (spatial regression) against the separate fMRI data sets, resulting in matrices describing temporal dynamics for each component and participant, and (ii) using these time-course matrices in a linear model fit (temporal regression) against the associated fMRI data set to estimate subject-specific spatial maps. For each patient the individual lesion site was masked so that BOLD signal variance in these voxels did not contribute to this second step.

We focused on RSNs that were structurally most affected by the lesions. We calculated the degree of spatial overlap between individual patients’ lesions and the RSN normalised for the total spatial extent of each RSN. We calculated the total spatial overlap across all RSNs (excluding a component spatially contiguous with the ventricles) and compared between the two lesion groups with independent samples t-tests. For the analysis we explicitly focused on the default mode network and RSNs identified by Beckmann et al. (2005). We identified all eight RSNs reported by Beckmann: 1. the medial visual network, 2. the lateral visual network (which had divided into two subcomponents), 3. auditory network, 4. sensory motor network (which had also divided into two components), 5. visuo-motor network, 6. executive Control network and the 7. left and 8. right dorsal visual stream. We also confidently identified the default mode network, which was deconstructed into an anterior and posterior component. From these 12 RSNs we determined the top three spatially coincident RSNs in each lesion group and compared the degree of lesion overlap of each individual across the two patient groups with independent samples t-tests. On these top spatially contiguous RSNs we performed the third and final step of the dual regression pipeline. The RSN component maps were concatenated across all participants into single 4D files (1 per original ICA map, with the fourth dimension being subject identification) before being subjected to non-parametric permutation testing (randomise *n* = 10,000). Again cluster-based thresholding (clusterm c = 2.3, p < 0.05) (Nichols & Holmes, 2002) was calculated over a small volume correction GM mask. This mask excluded lesioned voxels and was restricted to voxels that fulfilled either of the following criteria [1] the spatial extent of the RSN of interest or [2] the spatial overlap between VBM lesion effects and sRS network in healthy Controls (see *Supplementary Methods: Seeded resting state*). As such, even though the GLM was the same, Controls were compared to patient groups in two separate GLM analysis with different GM inclusion masks. The GLM included group as a factor, as well as the confound regressors of age, sex, handedness and number of years in education. The GLM included two contrasts; Controls > Lesion group and Lesion group > Controls. For illustration, all effects were then up-sampled to 2 mm^3^ resolution.

## Results

### Participant characteristics

Lesions were traced from the available CT or MRI onto the standard Montreal Neurological Institute (MNI) brain using MRIcro software. In two cases, lesions were labelled manually on a T1-weighted MPRAGE image prior to registration to the MNI brain. All lesions were overlaid to define the cluster of maximum overlap (lesion overlap image files are availale in supplementary material). In the 7 patients with SFG damage, the centre of gravity of this cluster fell within dorsomedial WM (−15, 11, 50). The overlap in all 6 rIFG patients was also in WM (28,31,11). Figure 2 shows the overlap image of lesion tracings for each group. We calculated the maximal number of patients with voxels damaged within a lesioned area. Despite variability in lesion location and extent, voxels in a cluster sized 3704 mm^3^ (centered on MNI coordinates of −15, 11, 50) were damaged in all SFG patients. For the rIFG group all lesions overlapped in a 560 mm^3^ WM cluster (centered on MNI coordinates of 28, 31, 11). The two lesion groups did not differ in lesion volume (t_11_ = 1.67, p = 0.124).

**Figure 1.**
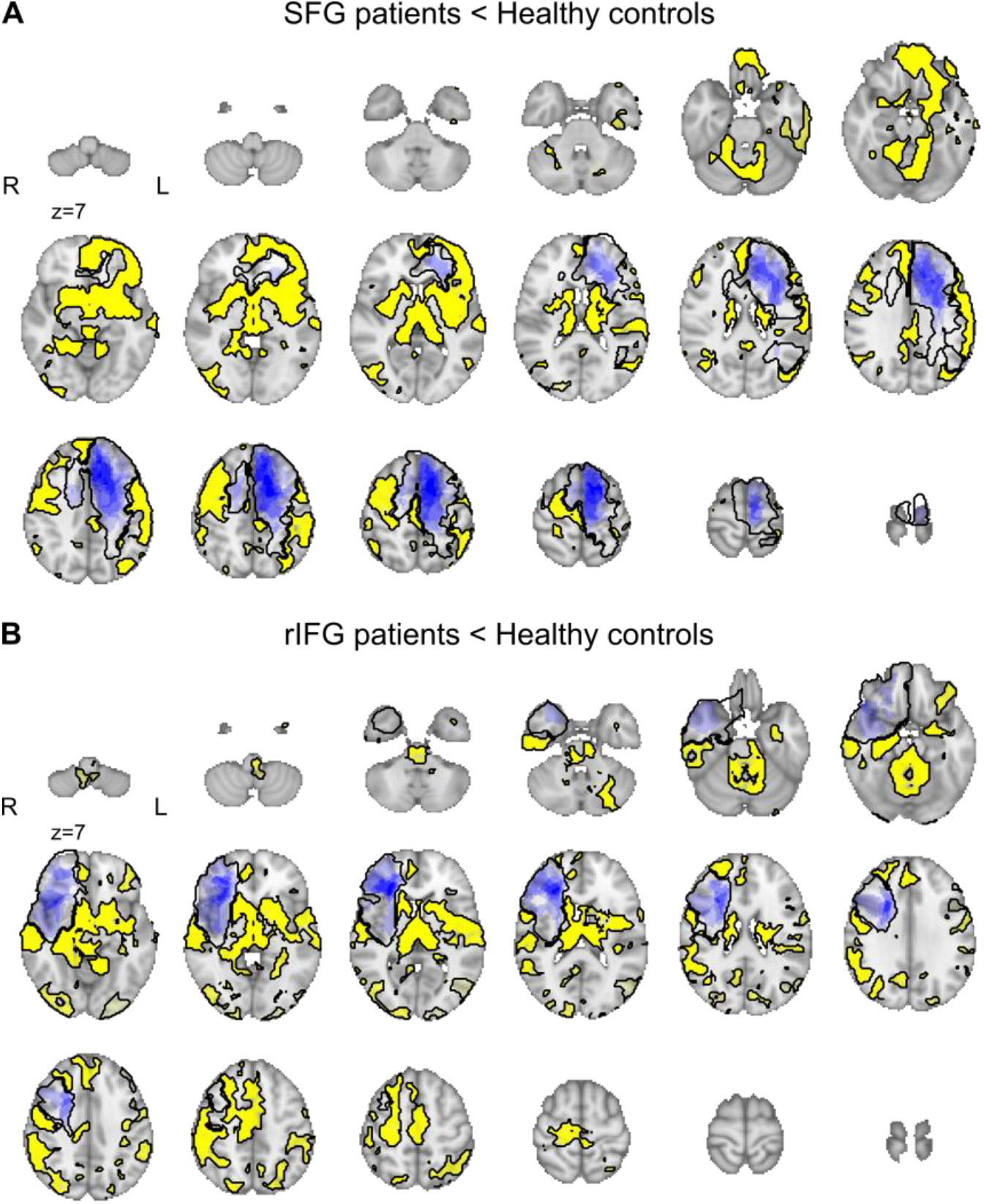
Lesion overlap (blue; intensity reflects number of participants) and VBM lesion effects (yellow) for Controls > SFG (A) and Cont>rIFG (B). Brain slices increase by internals of 8 mms from the most ventral slice of z = 7.

**Figure 2.**
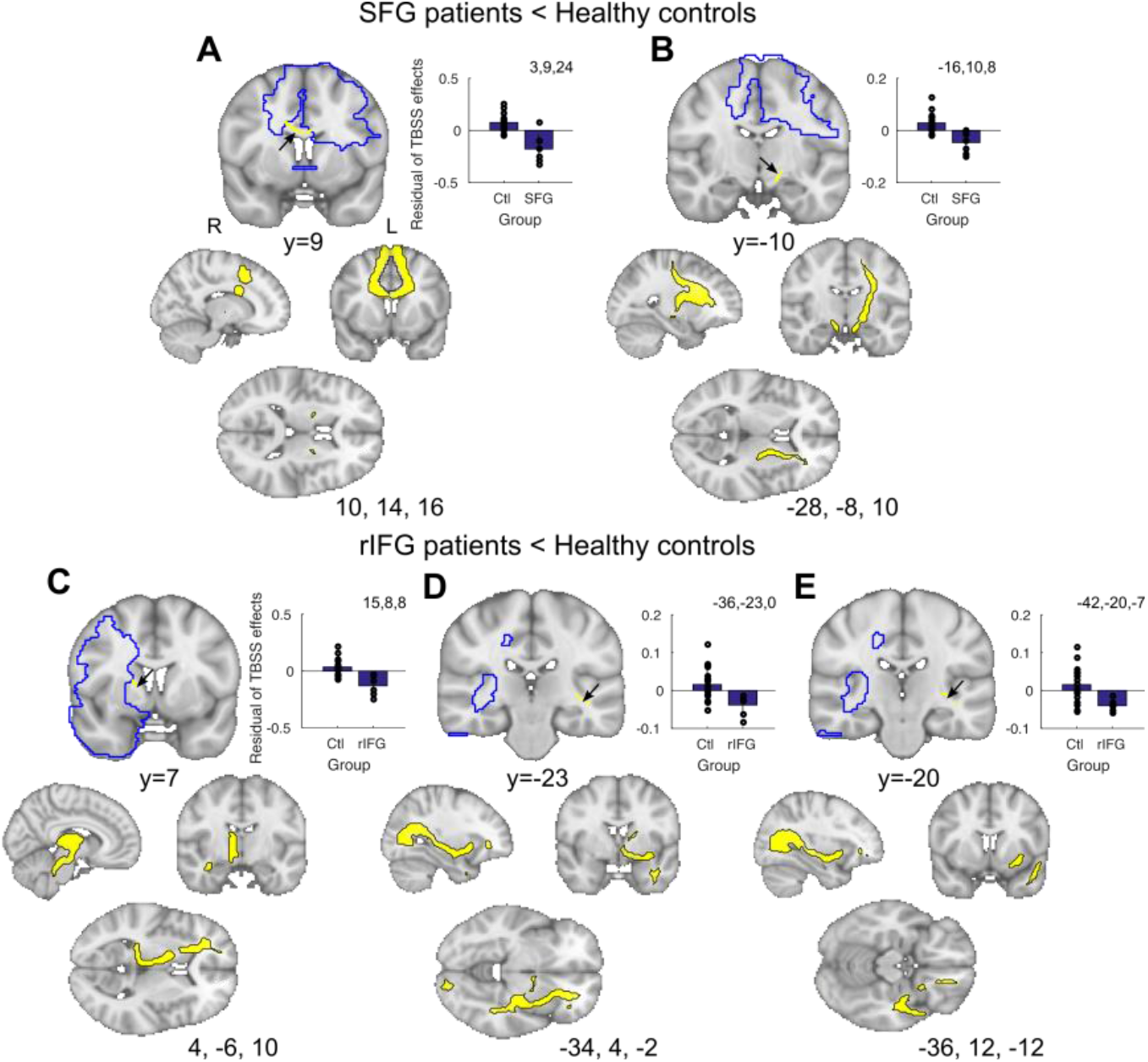
Effects of superior frontal gyrus (SFG) and right inferior frontal gyrus (rIFG) lesions on fractional anisotropy (FA). TBSS results show reduced FA in the corpus callosum and contralesional cerebral peduncle after SFG lesions (arrows in A and B), and reduced FA in bilateral internal capsule and contralesional ILF/IFOF after rIFG lesions (only illustrating right hemisphere effects; green arrows in C, D, E). Residual TBSS WM effects extracted from clusters identified in the group level lesion contrasts. Lower panels: tracts were visualised with probabilistic tractography seeded from the TBSS effects (depicted in yellow).

Demographic information and neuropsychological screening test results are provided in Tables 1 and 2, respectively. Controls were similar to both patient groups in age (p values => 0.236) and education (p values => 0.291). Further, the patient groups did not differ on the Beck Depression Inventory (p = 0.432), estimated IQ (p = 0.500), Animal Fluency (p = 0.666), F-A-S Fluency (p = 0.851) or backwards digit span (p = 0.576).

### Defining the lesion site

As the maximal site of lesion overlap for both groups was in WM (Figure 1), we used connectivity analyses to determine the cortical structures most likely affected by these lesions. Using probabilistic tractography seeded from the WM lesion overlap coordinates in healthy Control participants we estimated connectivity to cortical and subcortical GM anatomical targets. This analysis is the first step in a series that defines the network and network parameters of the SFG and rIFG in healthy Controls (described below). Analysis details are provided in Supplementary Methods: *Network definition and parameter measures in healthy Controls*. Results, shown in Supplementary figure S4A suggest that the SFG WM seed is most structurally connected with the SFG, but also to a lesser extent with the MFG and cingulate regions. The IFG WM seed is most connected with the IFG (pars triangularis) but also closely coupled with frontal polar cortex.

### Extended differences in structural morphology beyond the lesion sites

We performed a voxel-based morphology (VBM) analysis to identify the extended GM network altered by SFG or rIFG lesions relative to Controls. As illustrated in Figure 1, GM changes extended well beyond the lesion site. Our results show that SFG patients have reduced GM relative to Controls in superior frontal gyrus, caudate, thalamus and frontopolar cortex and insula (full VBM contrast maps are availale in supplementary material).

RIFG patients showed reduced GM relative to Controls in a number of brain regions that includes temporal cortex, temporoparietal cortex, middle, and inferior, frontal gyrus, caudate, thalamus, brainstem, cerebellum and orbitofrontal cortex (see VBM contrast maps). The total number of voxels significantly different relative to Controls were similar in the two groups (SFG = 48,211 and rIFG = 49,479). No clusters in which GM was larger in patients compared to Controls survived correction for multiple comparisons.

### Connectivity of the SFG and IFG in healthy Controls

The aim of the next analyses was to confirm that the network identified by the VBM analysis is what we might expect from transneuronal degeneration from the lesion site. We performed two connectivity analyses in healthy Controls using DMRI and resting state data to define the normal connectivity of each lesion site. Detailed methods are described in the Supplementary material.

#### Healthy WM network

First, we seeded probabilistic tractography in WM within the two lesion sites, and a control site in lateral orbitfrontal gyrus (LOFG; see *Supplementary Methods: Probabilistic tractography*). Tractograms and WM projections are illustrated and described in Supplementary results and shown in figure S1A-C. We quantified the degree of overlap between the DMRI and VBM lesion-derived networks by using the VBM effects as classification masks and taking the median connectivity value between each WM ROI seed and all significant VBM voxels in each participant. As expected, tracts seeded from the SFG WM seed reached the SFG VBM lesion affected regions more compared to a control LOFG WM seed (Supplementary figure S1D, SFG v LOFG t_16_ = 6.39, p < 0.001). Similarly, as predicted, tracts seeded from the rIFG are more likely to connect with IFG VMB lesion effects than the control LOFG WM seed (Supplementary figure S1E, IFG v LOFG t_16_ = 10.75, p < 0.001).

#### Healthy functional connectivity (FC) network

Second, again in healthy Controls, we seeded separate resting state analyses in GM coordinates of the SFG and rIFG lesion sites (see *Supplementary Methods: Seeded resting state*), and the LOFG as a control site. We identified regions that were significantly coupled to the SFG, rIFG and LOFG seeds and compared the spatial topography of these seeded resting state (sRS) networks with the topography of the VBM lesion effects. SRS and VBM overlap for the two FC analyses is described and illustrated in supplementary results and shown in figure S2. We quantified the degree of overlap between sRS and VBM lesion derived networks, using the Harvard Oxford parcellation atlas (See *Supplementary Material: Appendix Harvard Oxford Atlas*) to identify the proportion of ROIs (excluding those affected by the lesion) in which significant sRS effects for each network overlapped with significant VBM effects (*Supplementary Table S1*). As expected, as a proportion of size of the sRS network, both sRS SFG and sRS IFG network overlapped with their respective VBM networks to a greater degree than the sRS LOFG control network. In the SFG case over 100% more VBM ROIs overlapped with a SFG sRS network than LOFG sRS network. In the rIFG case, 29% more VBM ROIs overlapped with a rIFG sRS network than LOFG sRS network. These independent analyses in healthy Controls provide evidence that lesions to these two regions likely result in distributed changes at distal GM regions identified in the VBM analyses. The WM tracts and sRS networks defined in healthy Controls were used to statistically constrain later analyses in the lesion groups.

### Limited differences in structural connectivity of WM tracts after frontal lobe lesions

Next, we analysed the impact of lesions to the structural integrity of WM by measuring differences in fractional anisotropy. We performed a TBSS analysis to identify the WM network altered by SFG or rIFG lesions relative to Controls. Note, the analyses were constrained to the tracts identified using probabilistic tractography in healthy Controls as emanating from the lesion site coordinates. Unlike GM lesion effects, WM effects were relatively limited and often localised close to the lesion site (Fig 2). Our results show that SFG patients show reduced WM relative to Controls in the corpus callosum (629 voxels, MNI: 5, 15, 19) and contra-lesional cerebral peduncle (74 voxels, MNI: −13, −9, −8).

By contrast, rIFG patients show reduced WM FA relative to Controls in bilateral internal capsule (which included the retrolenticular portion and Extreme Capsule (EmC) in the contralesional hemisphere 350 voxels, MNI: 19,10,9 and 49 voxels, MNI: −32,-23,0), as well as contralesional ILF/IFOF (Inferior longitudinal fasciculus/inferior fronto-occipital fasciculus; 41 voxel, MNI: −40,-22,-8). Tracts were visualised with probabilistic tractography seeded from the TBSS effects (lower panels) and identity confirmed with the Xtract WM atlas. No WM clusters survived correction for multiple comparisons in a contrast examining greater FA in Lesioned Grops compared to Controls.

### Limited differences in resting state networks after frontal lobe lesions

The final key analysis investigated functional differences within RSNs between Lesion Groups and Controls using dual regression methods (Fig 3A). We hypothesised that key networks putatively affected in the SFG and rIFG groups should be those in which lesions were most coincident with the spatial topography of RSNs. While lesions in the SFG group overlapped with all RSNs to a greater degree than rIFG patients (t_11_ = 3.49, p = 0.005) we focused on three networks that overlap most with each lesion group (Fig 3B-E). For the SFG group, these were executive control network (ECN), posterior sensorimotor network (pSMN), and the anterior component of the default mode network (aDMN). For the rIFG group, the RSNs were right dorsal attention network (rDAS), ECN and aDMN. We hypothesised that these RSNs would show altered functional connectivity in the respective lesion groups relative to Controls. We compared the relative degree of overlap between the two lesion groups and each of the four RSNs. As expected given our methods of selection the pSMN most overlapped with the lesions of those in the SFG group (t_11_ = 3.31, p = 0.007) and the rDAS (t_11_ = −4.29, p = 0.001) overlapped most with the lesions in the rIFG group. However, the relative overlap between lesions the lesion groups and the aDMN or ECN did not differ (aDMN: t_11_ = 0.96, p = 0.356; ECN: t_11_ = 0.97, p = 0.353) with the two groups equally spatially overlapping these two networks.

**Figure 3.**
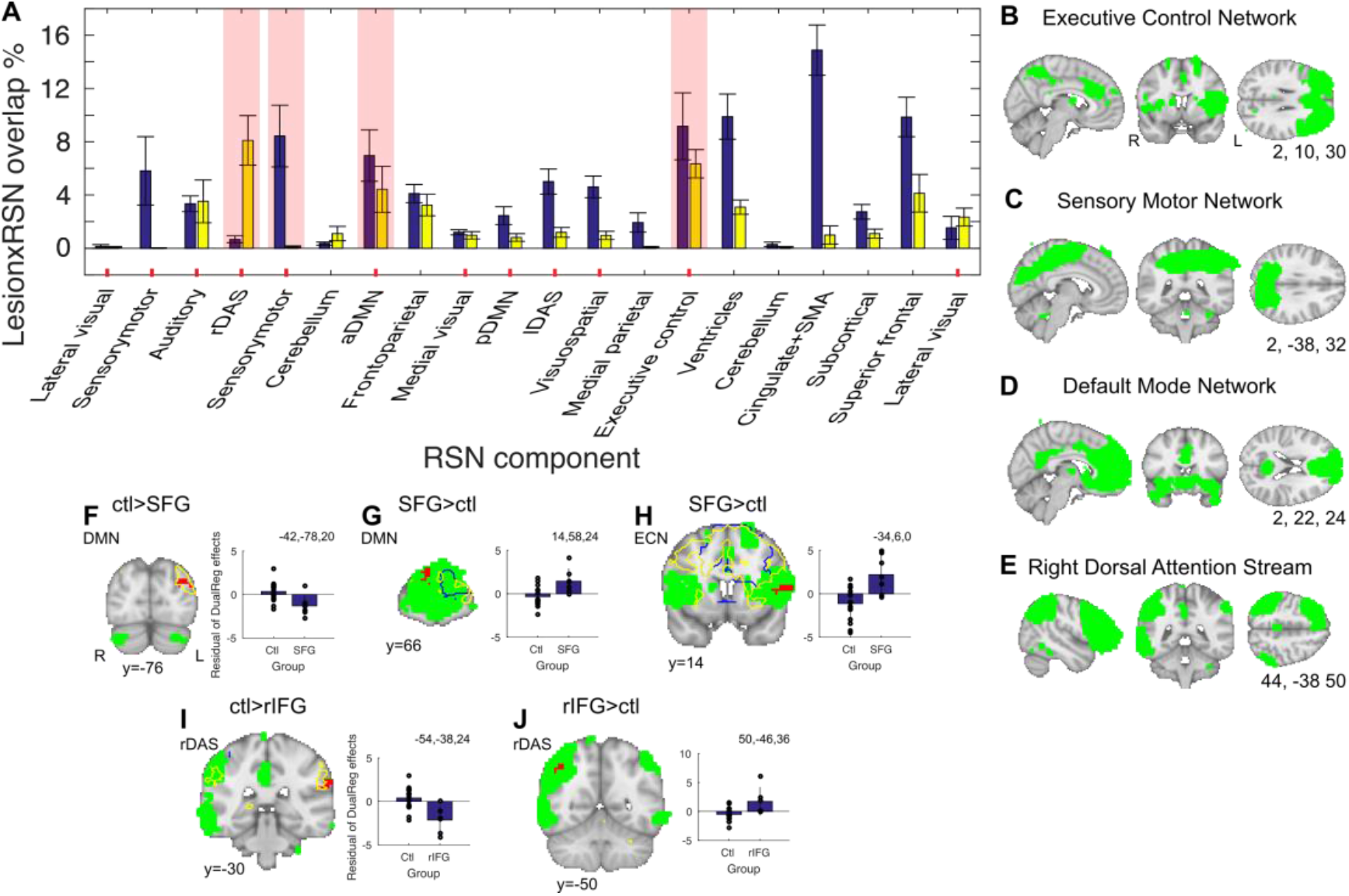
Resting state networks (RSNs) functional connectivity. (A) Top three RSNs for each Lesion group that overlap with the SFG (blue) and rIFG (yellow) patient lesion sites. The superior frontal gyrus lesions overlap most with the Executive Control Network (ECN), posterior component of the Sensorymotor Network (pSMN) and anterior Default Mode Network (aDMN). The right inferior frontal gyrus (rIFG) overlap most with the right Dorsal Attention Stream (rDAS), the ECN and the aDMN. Red ticks indicate the 12 RSNs identified either from Beckmann or as the DMN. Red highlights indicate the top 3 spatial maps of each Lesion group of those 12 identified RSNs. (B-E) Spatial maps of the RSNs that most overlap with the patient groups lesions (blue lesion overlaps shown in Figure 1). From top to bottom: executive control network, sensory motor network, anterior default mode network, and right dorsal attention stream. (F-J) SFG patients show reduced functional connectivity (FC) between left lateral occipital cortex connectivity with aDMN (F), increased FC between right rostrolateral frontal polar cortex connectivity and aDMN (G), and increased FC between left Inferior frontal gyrus / frontal operculum connectivity and ECN (H). rIFG patients show decreased FC between rDAS network and left supramaginal gyrus cortex connectivity (I), and increased FC between rDAS network and right angular gyrus cortex (J). Total lesion outline represented in blue, outline of overlap between VBM GM effects and seeded RSN in yellow, ICA network in green, dual regression effects in red. Residual dual regression effects extracted from clusters identified in the group level lesion contrasts.

Given the wide extent of the GM changes observed in both lesion groups, we found surprising conservation of functional networks. However, there were some local differences in connectivity within some networks. First, comparisons between SFG patients and Controls revealed that the ipsilesional left lateral occipital cortex showed reduced functional coupling with the aDMN in SFG patients (Fig 3F, p = 0.006). In the reverse contrast, there was more functional coupling between the aDMN and the contralesional right rostrolateral frontopolar cortex in SFG patients (Fig 3G, p = 0.013). Finally, a cluster at the intersection of the ipsilesional left inferior frontal gyrus, frontal operculum and insular cortex was more coupled to the ECN in patients than Controls (Fig 3H, p = 0.006). No other significant differences in functional connectivity between SFG patients and Controls were identified (see Supplementary table S2 for strongest clusters peaks).

By contrast lesions to the rIFG significantly affected coupling within the rDAS, with decreased functional connectivity in the patients, relative to Controls, between this network and contralesional left supramaginal gyrus (Fig 3I, p = 0.025) and increased coupling with the ipsilesional right angular gyrus (Fig 3J, p = 0.040). No regional clusters reached significance between rIFG patients and Controls for the other RSNs tested (see Supplementary table S2 for strongest clusters peaks).

### Corroboration of functional connectivity using the Neurosynth database

To independently corroborate our functional connectivity results and to understand the wider connectivity of the lesion site in relation to the published literature, we seeded the SFG (−18, 10, 50) and rIFG (32, 30, 6) lesion sites in the Neurosynth database (see *Supplementary Methods and Supplementary Figure S3*). Results suggest the SFG lesion site, as well as being connected to its contralateral homologue, functionally connects to only two clusters in the brain, located in extrastriate lateral occipital cortex (Fig S3A and B, LOC; cluster = 62, peak z = 0.227, MNI: −32 −82 36; cluster = 49, peak z = 0.223, MNI: −6 −68 54); one of which lies 10 mm medially and dorsally to the cluster found here to differentially connect with the aDMN. The Neurosynth database shows that the SFG and this LOC region tend to co-activate in motor imagery (z = 3.79) and simulation (z = 4.95) studies.

By contrast, the rIFG, in addition to its contralateral counterpart, is functionally connected to the dorsomedial PFC (Fig S3C cluster = 1296, peak z = 0.328, MNI: 4 14 50), bilateral supramarginal gyrus (Fig S3D cluster = 688, peak z = 0.282, MNI: 62 −32 36; cluster = 185, peak z = 0.231, MNI: −60 −36 28) and bilateral superior frontal gyrus (Fig S3E cluster = 440, peak z =0.275, MNI: 36 46 32; cluster = 127, peak z = 0.23, MNI: −36 44 36). Co-activation association analysis suggest that these regions tend to co-activate in pitch perception (z = 4.57), language (z = 3.81) and pain (z = 3.55) studies.

### Network parameters in healthy Controls

To independently test the model prediction that midline lesions would have more widespread effects than lesions to lateral regions of cortex (Alstott *et al*., 2009) we quantified the connectivity of the SFG and IFG using WM and resting state fMRI in healthy Control participants using common summary measures of node connectivity (Alstott *et al*., 2009; Hwang *et al*., 2013), see *Supplementary Methods: Network definition and parameter measures in healthy Controls*. For WM connectivity we calculated the total number of cortical and subcortical GM target ROIs reached by probabilistic tractography tracts seeded from the two lesion overlap sites (degree of connectivity). The results suggested that the IFG WM was more highly connected to the rest of the brain than the SFG WM site (Supplementary Figure S4B, degree t_16_ = −3.12, p = 0.006). By contrast, in resting state data a partial correlation analysis of BOLD timeseries extracted from cortical and subcortical GM anatomical targets identified the number of significant paired regions (degree of connectivity) and the sum of the absolute partial correlation coefficient (strength of connectivity) for the SFG and rIFG seeded regions (Supplementary Figure S4C). Contrary to WM network metrics, both degree and strength metrics suggested that the SFG had greater network connectedness than the IFG (Degree t_17_ =4.31, p < 0.001; Strength t_17_ = 4.00, p < 0.001, Supplementary figure S4D and S4E respectively).

### Control analyses

We performed a number of control analyses to rule out involvement of confound variables (see *Supplementary Methods*). Lesion groups showed no differences in whole brain parameters of intracranial volume (ts =< −1.97, ps=>0.061) or frame displacement (ts =< 0.34, ps => 0.732) compared to Controls, arguing that these confounds are unlikely to explain the observed effects.

## Discussion

This study looked for converging evidence of the effects of circumscribed damage on functional and structural networks. For practical reasons, in this proof-of-principle study, we focused on damage to two regions of the frontal lobes, the superior and inferior frontal gyrus, both putatively key nodes in functional networks important in higher-order cognition (Beckmann *et al*., 2005; Aron *et al*., 2014; Rae *et al*., 2015). GM differences were identified between patients and Controls as a first step and we used complementary, orthogonal network analysis in healthy participants to define the normal networks. We then compared structural and functional connectivity between patients and Controls to identify network-wide differences in WM or GM identified as networked with the lesion site, via orthogonal analyses. We also tested predictions of the effects of lesions on brain organization derived from anatomically-informed models of large-scale functional and structural connectivity (Alstott *et al*., 2009), aiming to resolve discrepancies in the existing lesion literature (Nomura *et al*., 2010; Eldaief *et al*., 2017). The results suggest that chronic focal damage had effects on distant parts of the brain in both frontal lobe groups, with the extent differing substantially depending on how the network was probed.

### Lesion impact on brain networks networks depend on modality and location

Our primary analysis examined the extent to which lesions affected distant GM, WM and RSNs. Alstott et al. (2009) made simulated lesions in regions overlapping with our SFG and IFG lesions in a computational model and examined changes in resting state and diffusion-derived networks. This yielded the prediction that functional connections would be more affected by lesions than structural connections. Our experimental results however do not support this hypothesis. Instead, we found relative preservation of both of these functional and structural network metrics in the lesion groups (discussed below). By contrast, GM appears most sensitive to the impact of lesions: we found evidence of widespread differentiable effects in GM volume far beyond both local lesion sites, relative to healthy Controls. SFG damage was associated with large disruptions in GM volume in an extended superior-fronto-cortico-thalamic network and rIFG damage was associated with reduced GM in a more inferior fronto-cortico-thalamic network.

Across imaging modalities, we also tested the model prediction that midline lesions would have more widespread effects than lesions to lateral regions of cortex (Alstott *et al*., 2009). The findings provide only limited support for this claim. Our analysis of network parameters in healthy brains suggested that while the SFG is a more widenly-interconnected node when measured with functional connectivity, rIFG is more connected to other brain regions through WM tracts. By contrast, analysis of the lesion data suggest that structural network metrics (i.e. GM volume and fractional anisotropy) appear to be affected to a comparable degree by midline and lateral lesions. But, in line with the connectivity analysis in healthy Controls, midline lesions did have greater impact on functional connectivity than lateral lesions. Despite no overall difference in lesion extent, SFG lesions overlapped to a greater degree with all RSN compared to rIFG lesions. Further, while SFG and rIFG lesion spatially overlap with the DMN and ECN to the same degree, only the SFG lesion group showed changes within these two networks.

These mixed findings are representative of the current literature. While cortical structures on the midline are often predicted by network estimates derived from healthy brain networks, as well as macaque and human lesion models, to have the greatest potential to cause widespread brain functional network disruption (Honey & Sporns, 2008; Alstott *et al*., 2009) there is little direct evidence from lesion patients to support this. Consistent with our findings, midline mPFC lesions do not alter intrinsic functional connectivity among undamaged DMN nodes (Eldaief *et al*., 2017). Instead, like the SFG RSN effects reported here, network-specific changes manifest as weaker correlations between whole brain RSNs.

### A non-linear relationship between structural and functional networks

Overall, this multimodal neuroimaging approach in the same participants provided little evidence that distal GM volume loss is underpinned by degeneration of intervening WM tracts. While trans-neuronal degeneration can be caused by a loss of signal input (Heimer & Kalil, 1978) and relationships between GM and WM degeneration have been observed in people with Alzheimer’s disease (Jang *et al*., 2017), our study suggests this is not always the case, at least as measured by current methods. Only the corpus callosum and internal capsule (predominantly the cerebral peduncle and corticospinal pathways) showed reduced structural integrity in SFG patients relative to Controls, while rIFG lesions only affected WM metrics in the internal capsule (with tractography suggesting the affected fibres project to thalamic frontal tracts and EmC) as well as projecting through ILF & IFOF, UF tracts) (Schmahmann & Pandya, 2006). This pattern of effects may indicate that despite lesion heterogeneity, WM involved in motor control is most vulnerable at the chronic lesion stage. The difference measures may therefore show effects at different timesscales (Berthier *et al*., 2011). Alternatively, these large, well-defined tracts may be most likely to show detectable effects, for technical reasons.

We also found little evidence that GM degeneration affects functional connectivity. Although some recent work has linked RSNs and GM metrics in healthy young people (Hunt *et al*., 2016) and people with neurological conditions, such as Parkinson’s Disease (Lucas-Jimenez *et al*., 2016), our results suggest that chronic focal lesion effects on distant GM regions are only accompanied by small-scale changes in functional connectivity within well-established RSNs. We identified both reduced and increased (potentially compensatory) activity in patient groups relative to Controls. For example, in SFG patients we found that lateral occipital cortex connectivity with the aDMN was significantly reduced compared to Controls, while increased connectivity was found between aDMN and rostral frontopolar cortex, as well as between the IFG and the ECN. A similar pattern was seen in the rIFG patients patients, with reduced functional coupling between rDAS and contralesional supramaginal gyrus, but increased coupling with the ipsilesional angular gyrus. This overall pattern of network preservation begs the question: How can functional networks be so globally robust to lesions which affect network nodes? This question is further complicated by evidence that nodes have unequal status (van den Heuvel & Sporns, 2013). One possibility is that the RSNs studied here (at least) emerge due to common (perhaps subcortical) drivers rather than cortico-cortical structural connections.

### Limitations and future directions

Analysis of multimodal imaging comes with a number of challenges and limitations. For example, the degrees of freedom within analysis methodology could lead to false positives if enough analyses are run. To reduce this possibility, we constrained our analyses to the most widely used and validated platforms within the FSL toolbox. While we do not compare directly across patient groups or data types because of limited sample size, future studies with more power may also subject this type of data to exploratory analysis tools designed to automatically find patterns of effects consistent across imaging modalities (Groves *et al*., 2011). Furthermore, future studies should include task-related FC measures as well as resting state FC, as dynamic diaschisis has been described following injury with differences only apparent in certain contexts (Price *et al*., 2001). Our study also does not probe the potential relationship between functional/structural connectivity and behaviour (Siegel *et al*., 2018). Finally, patient data itself provides additional challenges. For example, some lesions may pose problems for standard segmentation and registration software. To minimize this possibility, we meticulously checked segmentation and registration images and confirmed no outliers drove the group differences. Further, in the absence of longitudinal measures it is challenging to ascertain whether the findings we observed are secondary to a pathological state (diaschisis) or arise as compensatory mechanisms during recovery (Carrera & Tononi, 2014). Our study does not test the effect of lesion chronicity on multimodal network connectivity. Longitudinal data accompanied by behavioral measures will be needed to address these important questions (see Fornito *et al*., 2015 for discussion).

## Conclusions

We applied multimodal structural and functional neuroimaging to examine the effects of focal damage to two regions of the frontal lobes on structural and functional networks. Extensive differences in GM volume were evident beyond the lesion site, relative to Controls, but WM paths and functional networks were largely conserved. Some limited functional connectivity differences were found in resting state networks thought to underpin higher-order cognitive processes, but WM differences were only detected in cortico-motor pathways. The findings shed light on the potential neural substrates of widely studied RSNs, showing unexpected discordance between different structural and functional measures, and also can be used to refine existing computational models of such brain networks.

## Data Availability

Data not available due to ethical restrictions

## Acknowledgements

The authors would like to thank Saad Jbabdi for his support with the analysis pipelines, Arlene Berg for assisting in participant recruitment and the volunteers who participated in the research. The work was supported by the Canadian Institutes of Health Research. The work of R.B.M. is supported by the Biotechnology and Biological Sciences Research Council UK [BB/N019814/1] and the Netherlands Organization for Scientific Research [452-13-015]. The Wellcome Centre for Integrative Neuroimaging is supported by core funding from the Wellcome Trust [203129/Z/16/Z]. M.R.G. has received research funding from the Canadian Institutes of Health Research, National Institutes of Health, Sidney Baer Foundation and Fonds de Recherche en Santé du Québec (Chercheur-Boursier Award). LKF acknowledges support from McGill University’s Healthy Brains for Healthy Lives Canada First Research Excellence Fund.

