## Supplementary Methods for "Characterisation of multimodal network organisation after focal prefrontal lesions in humans"

To quantify the VBM effects and inform our interpretation, we examined connectivity of the two lesion sites in the healthy Control group, as well as a non-lesioned control site. We therefore conducted two additional connectivity analyses in the healthy control sample: probabilistic tractography and seeded resting state.

*Probabilistic tractography: From lesion site to VBM effects*

In healthy control subjects, we defined the structural connectivity between the lesioned brain areas and the VBM lesion effects. The lesion overlap sites (SFG MNI -15, 11, 50 and rIFG MNI 28,31,11, see Results: Patient Characteristics for more detail) were used as WM ROI seeds ($216mm^3$), registered to individual subject space, in a probabilistic tractography analysis in healthy controls. Voxel-wise estimates of the fiber orientation distribution were calculated using Bedpostx, limited to estimating two fiber orientations at each voxel, because of the b value and number of gradient orientations in the diffusion data (Behrens, Berg et al. 2007 http://fsl.fmrib.ox.ac.uk/fsl/fslwiki/FDT). Probabilistic tractography was run for each subject, using a model accounting for multiple fiber orientations in each voxel. Five thousand sample streamlines were seeded from each voxel within each individual's seed mask. The tractography algorithm parameters used were a maximum of 2000 steps; step size of 0.5 mm and a curvature threshold of 0.2. Each streamline followed local orientations sampled from the posterior distribution given by BedpostX, as described previously. As a control site, and to reduce the possibility of biasing the results if one lesion site had a broader connectivity pattern than another, we also ran probabilistic tractography from the lateral occipital fusiform gyrus (LOFG, -14,-74,-12). The significant VBM effects acted as classification targets with probtrackx quantifying the connectivity values between the WM ROI seed mask (SFG, rIFG and LOFG) and the GM target mask (SFG lesion VBM effects and rIFG lesion VBM effects). The values in the resulting image file represent the number of samples seeded from that voxel reaching the relevant target mask voxels. We summed the connectivity values across GM seed voxels for each subject and normalised by the product of the size of the seed mask. We contrasted the degree of connectivity between each WM seed (SFG and IFG) and their respective lesion VBM effects against the same metrics derived from the control seed (ie [1] SFG WM seed –> SFG VBM vs LOFG WM seed –> SFG VBM and [2] IFG WM seed –> IFG VBM vs LOFG WM seed –> IFG VBM). Across subject differences in connectivity were quantified using paired-samples t tests.

Visitation maps referred to as tractograms were constructed for each individual from the raw output connectivity distribution (ie unconstrained by VBM effects). These connectivity distribution values were log transformed, normalized by dividing by the maximum tracts identified for each subject, thresholded at 0.8 and binarised (Mars, Foxley et al. 2015). We sought only the top 20% of tracts emanating from the seed mask. Finally, the tracts were summed across subjects, registered to MNI space and are illustrated thresholded at more than 50% of subjects. These tracts were used to constrain the lesion TBSS analyses described in the main manuscript, statistically acting as a small volume of interest.

*Seeded resting state: From lesion site to VBM effects*

In healthy control subjects, we defined the networks based on the functional connectivity of the two anatomical regions identified in the patient sample. A 216 $mm^3$ mask was drawn over the GM voxel closest to the centre of the lesion overlap. In the SFG group, the mask was drawn at MNI coordinates -18,10,50. According to the Harvard-Oxford cortical atlas this area of GM lies within the superior frontal gyrus, although closely flanked by middle frontal gyrus, paracingulate gyrus and juxtapositional cortex. Neubert and colleagues refer to this region as preSMA (Neubert, Mars et al. 2015). Similarly, in the IFG group, we placed a 216$mm^3$ mask slightly lateral and ventral to the rIFG

WM coordinates in the nearest GM voxel at MNI coordinates 32,30,6. According to the Harvard-Oxford cortical atlas this area of GM is at the intersection of the inferior frontal gyrus, orbitofrontal cortex, insular cortex and frontopolar cortex. Neubert and colleagues refer to this region as area 45a (Neubert, Mars et al. 2014). As a control site, we placed a 216mm$^3$ mask in the lateral occipital fusiform gyrus (LOFG, MNI: -18,-74,-12). Cortex here was not damaged in either patient group. From these masks, the BOLD time series from all the healthy control subjects was extracted and used as an explanatory variable in a primary GLM analysis (FEAT). A nuisance regressor of the signal time series of the whole brain was also included in each primary-level analysis. Mixed effects analyses (FLAME 1 and 2) were applied to the whole brain group data in MNI space to generate statistical activation maps for each of the contrasts and to test for an effect of group. Group Z (Gaussianized t) statistic images were thresholded using clusters determined by $Z = 2.3$ and a corrected cluster extent significance threshold of $p = 0.05$. We then compared the spatial topography of the group-level seeded resting state networks with that of the VBM lesion effects (including lesioned voxels).

To quantify the degree of overlap we used the Harvard Oxford parcellation atlas. We indexed the seeded resting state (sRS) network by seeking those ROIs where group-wise significant sRS effects covered more than 25% of the ROI. Similarly, we indexed the VBM network by seeking those ROIs where significant VBM effects covered more than 25% of the ROI. We discounted any ROIs in which 50% of the voxels were damaged in any lesion patient. Overlap was then calculated as the proportion of ROIs in which the VBM effects overlapped with the sRS effects. This VBM x sRS overlap was also used, in part, to constrain the lesion dual regression analysis described in the main manuscript, statistically acting as a small volume of interest.

*Neurosynth analyses*

The Brain Genomics Superstruct Project within the neurosynth (http://neurosynth.org/) interface was used to create a map which represents resting-state functional connectivity analysis performed on 1,000 human subjects. We placed two seeds in GM voxels within the SFG (-18,10,50) and rIFG

(32,30,6) lesion site. Functional connectivity maps were thresholded at $z > 0.2$ and cluster extent above 100 voxels. Neurosynth meta-analytic coactivation maps were also generated from 6mm diameter spheres centred on the same voxel locations. Values represent z-scores quantifying the strength of association between the presence or absence of activation in each voxel in relation to the presence or absence of activation in the seed voxel. We report the highest-ranking z-scores for non-anatomy co-activation association terms. These represent the z-score value obtained at the current voxel in the "association test" meta-analysis map for the corresponding term.

*Network definition and parameter measures in healthy controls*

Using DMRI and resting state data from healthy controls, we defined the normal network of each lesion network and estimated network connectedness.

White matter network*:* In healthy controls, we defined the structural connectivity between the regions damaged in the lesion sample. The lesion overlap clusters identified above were registered to individual subject space and used as seeds ($216mm^3$) in a probabilistic tractography analysis in healthy controls using the same parameters described above.

Instead of the VBM effects acting as classification targets, in this analysis connectivity was estimated from each voxel within the two WM ROI seeds to every voxel in the brain. Each resulting connectivity weight was then categorized using the Harvard-Oxford cortical and subcortical atlases offering a partitioning of each hemisphere into 56 anatomically distinct regions– 48 cortical and 8 subcortical. For each subject, connectivity weights between each WM ROI seed voxel to any voxel in each of the 56 anatomical target ROIs were summed across seed voxels and the peak connectivity value within each target ROI was extracted and averaged across subjects. This analysis provided data-driven confirmation of our lesion grouping (see *Results: Defining the lesion site; Supplementary Figure 3A*). In all subjects, we also calculated the total number of target ROIs reached by any tract, regardless of connectivity weight, and compared the two WM ROI-seeded analyses with paired samples t-tests.

<u>Functional connectivity:</u> Network parameters were estimated from resting state data through a partial correlation analysis. Parcellations from Harvard Oxford cortical and subcortical atlas were used as anatomical ROIs. Anatomical ROI masks were registered to each healthy control's MRI scan and fMRI scan space in a step-wise manner, and the BOLD time series were extracted from each mask in each subject. We partialled out the confounding influence of the whole brain GM, WM, and cerebrospinal fluid (CSF) BOLD time courses by using the FSL general linear model (GLM) tool. We focused on coupling between the Harvard-Oxford SFG and IFG parcellations and each of the other 55 anatomical regions. The time series for pairs of regions were entered into four partial correlation analyses that each controlled for the correlation with the BOLD time series in all 55 other ROIs. Network parameters, degree, defined as the number of links connected to the node, and strength, defined as the sum of weights of outward links connected to the node, were calculated on a subject-wise basis as the sum of the significantly paired anatomical regions and the sum of the absolute partial correlation coefficient respectively. Paired t tests examined the between group difference in degree and strength. The resulting partial correlation values were then Fisher transformed and entered into a correlation and compared against zero in multiple Bonferroni-corrected t tests.

*Control analyses*

Whole brain parameters of intracranial volume and frame displacement (an index of movement) were measured as the total number of voxels in each subject's structural scan and the sum of frame-wise movement estimated from the resting state data during FSL pre-processing. Lesion groups were compared against controls using independent samples t-tests. Lesion groups were also compared on the number of years since lesion onset with independent samples t-tests.

**Supplementary Results**

*lesion overlaps*

The lesion overlap image files for the SFG and rIFG groups are available. They are titled SFGLesionOverlap.nii.gz and rIFGLesionOverlap.nii.gz respectively.

*Extended differences in structural morphology beyond the lesion sites*

Contrast maps of the VBM analyses for showing greater gray matter in healthy Controls than SFG lesion patients can be found in the supplementary image file titled VBM_Control>SFG_Corrp_tstat.nii.gz. Greater gray matter in healthy Controls relative to rIFG lesion patients can be found in found in VBM_Control>rIFG_Corrp_tstat.nii.gz.

*Connectivity of the SFG and IFG in healthy controls*

<u>Healthy WM network:</u> Supplementary figure 1 shows tractography seeded in the SFG courses through corpus callosum and corticospinal tracts. By contrast, rIFG seeded tractography utilises the internal capsule (Anterior thalamic radiation). The control LOFG seed is part of the inferior longitudinal fasciculus and uncinate fasciculus.

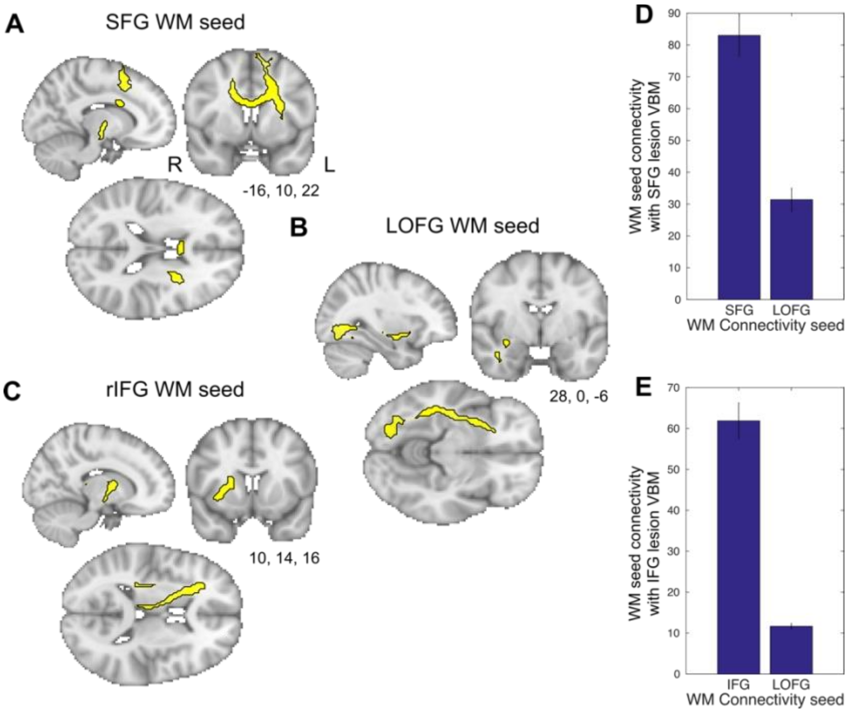

*Figure S1. Probabilistic tractography in healthy controls seeded from a 3mm$^3$ ROI at the coordinates of the SFG or rIFG lesion most commonly affected in the patient sample (A and B respectively). (C) Probabilistic tractography in healthy controls seeded from a 3mm$^3$ ROI in the control LOFG region. (D) Sum of probabilistic tractography connectivity weights in healthy controls between WM ROI seed and target VBM lesion effects comparing each WM lesion seed to the WM control seed.*

Healthy FC network: The results show that the SFG VBM lesion effects overlap with the functionally defined sRS network defined in the healthy brain (see supplementary results file sRSSFGcoord_Control_thresh_zstat.nii.gz for thresholded resting state contrast map). In addition to the dorsomedial cortex directly affected by the lesion, the functional connectivity network overlap extends beyond the lesion into other regions including contralesional SFG, medial frontal gyri, pre- and post-central sulcus, insula, as well as bilateral rostral frontal polar cortex and lateral occipital cortex (Fig S2A). The analysis also reveals overlap in the thalamus, putamen and hippocampus (see supplementary results file LesionVBMSFG_x_sRSSFG.nii.gz for full effects. Note to reproduce the overlap effects the VBM contrast maps e.g. VBM_Control>SFG_Corrp_tstat.nii.gz must be overlaid with the Lesion overlap images SFGLesionOverlap.nii.gz).

rIFG VBM lesion effects also overlap with the rIFG sRS defined network (Fig S2B; see sRSrIFGcoord_Control_thresh_zstat.nii.gz). As well as functional overlap within damaged cortex, the functional network extends and overlaps beyond the lesion site to contralesional IFG, bilateral paracingulate cortex and medial SFG/SMA, Planum Plare, supramaginal gyrus and superior temporal gyrus, as well as subcortically in the putamen (see results file LesionVBMrIFG_x_sRSrIFG.nii.gz for full effects). The control analysis, seeded in the LOFG (sRSLOFGcoord_Control_thresh_zstat.nii.gz), overlapped to a lesser degree with either VBM lesion effects (Fig. S2C,D, see results file LesionVBMSFG_x_sRSLOFG.nii.gz and LesionVBMrIFG_x_sRSLOFG.nii.gz for full effects).

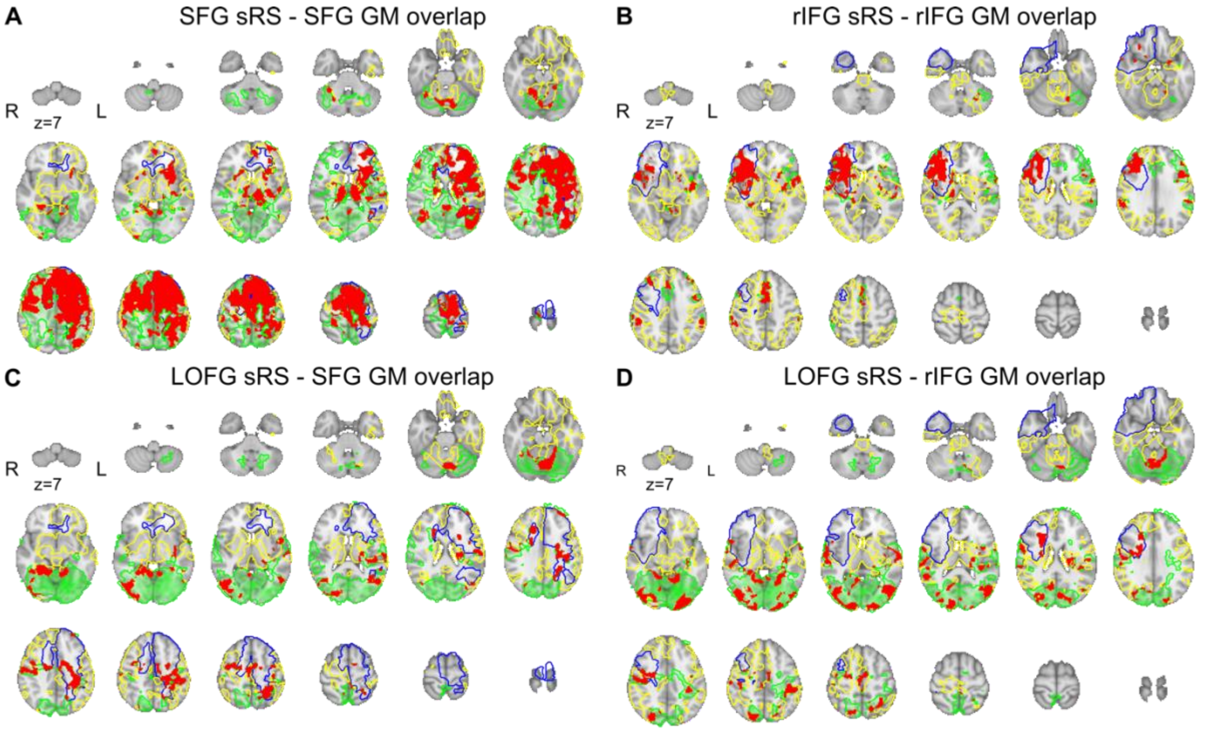


*Figure S2. Seeded resting state analysis in healthy Controls seeded from centre of gravity of the overlap shown of (A) seven SFG patients and (B) six rIFG patients. Seeded resting state network in green. SFG seed was moved laterally into the closest GM shown overlapped (red) with the SFG VBM lesions effects (yellow). rIFG seed was moved laterally and ventrally into the closest GM voxels shown overlapped with the rIFG VBM lesions effects (colour conventions are the same as the SFG seed). Lesion outline shown in blue. C and D show seeded resting state analysis in Controls seeded from the lateral occipital fusiform gyrus (green) shown overlapped in red with the (C) SFG VBM lesions effects and (D) rIFG VBM lesion effects. Brain slices increase in intervals of 8 mm from the most ventral slice of z = 7.*

*Table S1. Harvard Oxford ROIs identified in which VBM lesion effects overlap with the seeded resting state networks. This excludes any ROI with more than 50% lesion damage. Abbreviations in Appendix.*

| ***SFG VBM*** | | | | ***IFG VBM*** | | | |
|---|---|---|---|---|---|---|---|
| ***SFG sRS*** | | ***LOFG sRS*** | | ***IFG sRS*** | | ***LOFG sRS*** | |
| ***Region*** | ***num voxel overlap*** | ***Region*** | ***num voxel overlap*** | ***Region*** | ***num voxel overlap*** | ***Region*** | ***num voxel overlap*** |
| rMFG | 1700 | rLOCinf | 886 | lParaCG | 907 | rSTGpost | 561 |
| rPreC | 1995 | rLIN | 455 | lIC | 438 | rPostC | 917 |
| rParaCG | 742 | rTOFus | 316 | lSMGant | 421 | rSMGant | 474 |
| rPCG | 301 | lLOCsup | 1345 | lPOper | 222 | rLOCsup | 1557 |
| rLIN | 455 | lCOper | 412 | lPPolare | 292 | rLOCinf | 807 |
| rTHA | 1031 | lPOper | 169 | lPTemporale | 189 | rPTemporale | 291 |
| lFPole | 2996 | lPTemporale | 232 | lPUT | 749 | rOpole | 899 |
| lIC | 942 | | | lNAC | 56 | lLOCsup | 1358 |
| lIFGtriang | 419 | | | | | lLOCinf | 652 |
| lIFGoper | 345 | | | | | lCOper | 512 |

| | | | |
|---|---|---|---|
| lPreC | 1534 | lPOper | 222 |
| lPostC | 1254 | lPTemporale | 189 |
| lSMGant | 716 | lOpole | 755 |
| lSMGpost | 541 | | |
| lLOCsup | 1345 | | |
| lPCG | 565 | | |
| lFOper | 244 | | |
| lCOper | 412 | | |
| lHES | 235 | | |
| lTHA | 1138 | | |
| lCAU | 427 | | |
| lPUT | 799 | | |

*Limited differences in resting state networks after frontal lobe lesions*

*Table S2. Dual regression effects. Reporting p values of the strongest clusters, cluster size and corresponding peak MNI coordinates.*

| | | **Control>lesion** | | | **Lesion>control** | | |
|---|---|---|---|---|---|---|---|
| **Lesion** | **RSN** | **P value** | **Num voxels** | **MNI coord** | **P value** | **Num voxels** | **MNI coord** |
| **SFG** | **ECN** | 0.115 | 21 | - | 0.006* | 40 | -34, 6, 0 |
| | **pSMN** | 0.858 | 6 | - | 0.101 | 27 | - |
| | **DMN** | 0.006* | 36 | -42, -86, 20 | 0.013* | 10 | 30, 70, 12 |
| **rIFG** | **rDAS** | 0.025* | 25 | -54,-38, 24 | 0.04* | 33 | 50, -46, 36 |
| | **ECN** | 0.642 | 4 | - | 0.467 | 8 | - |
| | **DMN** | 0.806 | 5 | - | 0.167 | 20 | - |

* p < 0.05

*Corroboration of functional connectivity using the Neurosynth database*

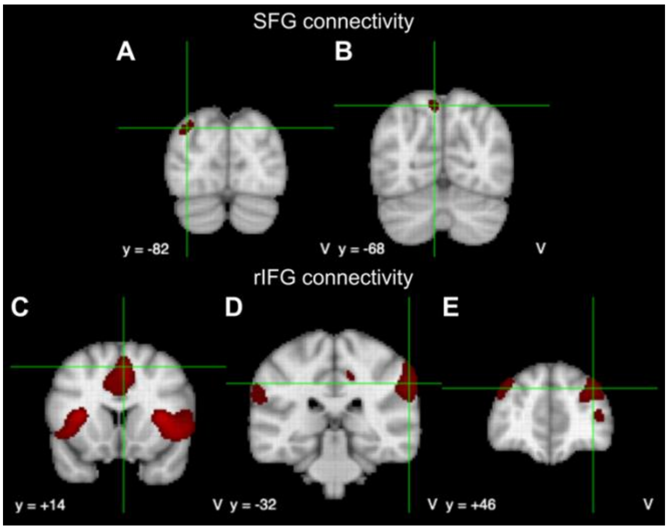

*Figure S3. Resting-state functional connectivity for a seed region in SFG (MNI -18, 10, 50) and rIFG (MNI 32, 30, 6) in the Neurosynth database of 1000 subjects. The SFG functionally connects to two clusters in extrastriate lateral occipital cortex (A and B). The rIFG functionally connects to the dorsomedial PFC (C), bilateral supramarginal gyrus (D) and bilateral superior frontal gyrus (E). Images thresholded between -0.2 and 0.2. Cross hairs placed on peak coordinates. Where bilateral regions were identified the coordinate is placed in the larger of the two clusters.*

*Network definition and parameters*

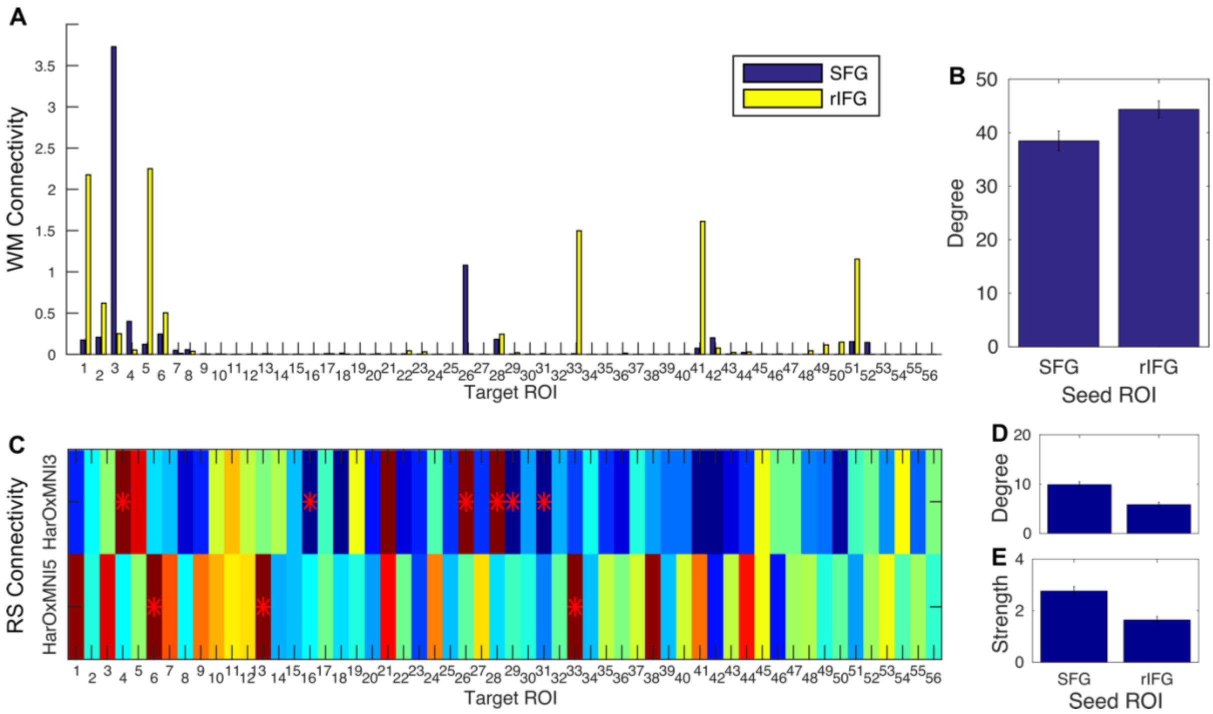


*Figure S4. Defining the lesion network in healthy controls. A. Mean of the maximum WM connectivity weight for each participant for each of the 56 Harvard Oxford parcellation target ROIs reached by WM ROI seeded tractography at the two lesion sites. B. Total number of target ROIs, regardless of weight, reached by WM ROI seeded tractography at the two lesion sites. C. Mean Fisher-transformed correlation coefficient representing the strength of BOLD coupling between each Harvard-Oxford parcellation ROI and the SFG and rIFG parcellation. D. Degree of resting state connectivity representing the mean total number of target ROIs in which BOLD activity is significantly coupled with the SFG and rIFG anatomical ROIs. E. Strength of connectivity representing the mean total number of target ROIs significantly paired with the SFG and rIFG anatomical ROIs.*

*Appendix Harvard Oxford Atlas*

*Table S3. 56 cortical and sub-cortical regions used to construct structural networks as defined by the Harvard-Oxford atlas. Abbreviations used: anterior division (AD); posterior division (PD); superior division (SD); inferior division (ID); temporooccipital part (TOP); cortex (ctx).*

| Label | Anatomical region | Label | Anatomical region |
|---|---|---|---|
| 1 | Frontal pole (FPole) | 29 | Cingulate gyrus AD (ACG) |
| 2 | Insular ctx (IC) | 30 | Cingulate gyrus PD (PCG) |
| 3 | Superior frontal gyrus (SFG) | 31 | Precuneous ctx (PCUN) |

| | | | |
|---|---|---|---|
| 4 | Middle frontal gyrus (MFG) | 32 | Cuneal cortex (CUN) |
| 5 | Inferior frontal gyrus, pars triangularis (IFGtriang) | 33 | Frontal orbital ctx (Forb) |
| 6 | Inferior frontal gyrus, pars opercularis (IFGoper) | 34 | Parahippocampal gyrus (PHIPant) |
| 7 | Precentral gyrus (PreC) | 35 | Parahippocampal gyrus (PHIPpost) |
| 8 | Temporal pole (TPole) | 36 | Lingual gyrus (LIN) |
| 9 | Superior temporal gyrus AD (STGant) | 37 | Temporal fusiform ctx (TFUSant) |
| 10 | Superior temporal gyrus PD (STGpost) | 38 | Temporal fusiform ctx (TFUSpost) |
| 11 | Middle temporal gyrus AD (MTGant) | 39 | Temporal occipital fusiform ctx (TOFus) |
| 12 | Middle temporal gyrus PD (MTGpost) | 40 | Occipital fusiform gyrus (OFus) |
| 13 | Middle temporal gyrus, TOP (MTGto) | 41 | Frontal operculum ctx (FOper) |
| 14 | Inferior temporal gyrus AD (ITGant) | 42 | Central opercular ctx (COper) |
| 15 | Inferior temporal gyrus PD (ITGpost) | 43 | Parietal operculum ctx (POper) |
| 16 | Inferior temporal gyrus, TOP (ITGto) | 44 | Planum polare (PPolare) |
| 17 | Postcentral gyrus (PostC) | 45 | Heschls gyrus (HES) |
| 18 | Superior parietal lobule (SPL) | 46 | Planum temporale (PTemporale) |
| 19 | Supramarginal gyrus AD (SMGant) | 47 | Supracalcarine ctx (SupraCAL) |
| 20 | Supramarginal gyrus PD (SMGpost) | 48 | Occipital pole (Opole) |
| 21 | Angular gyrus (ANG) | 49 | Thalamus (THA) |
| 22 | Lateral occipital ctx SD (LOCsup) | 50 | Caudate (CAU) |
| 23 | Lateral occipital ctx ID (LOCinf) | 51 | Putamen (PUT) |
| 24 | Intracalcarine ctx (IntraCAL) | 52 | Pallidum (PAL) |
| 25 | Frontal medial ctx (FMC) | 53 | Brain stem (BS) |
| 26 | Juxtapositional ctx (SMA) | 54 | Hippocampus (HIP) |
| 27 | Subcallosal ctx (subcallosal) | 55 | Amygdala (AMYG) |
| 28 | Paracingulate gyrus (ParaCG) | 56 | Accumbens (NAC) |
